# *Helicobacter pylori* seropositivity associates with hyperglycemia, but not obesity in Danish children and adolescents

**DOI:** 10.1101/2024.02.09.24302062

**Authors:** Sigri Kløve, Sara E. Stinson, Fie O. Romme, Julia Butt, Katrine B. Graversen, Morten A. V. Lund, Cilius E. Fonvig, Tim Waterboer, Guillermo I. Perez-Perez, Torben Hansen, Jens-Christian Holm, Sandra B. Andersen

**Author notes:** Correspondence: Sigri Kløve, Sandra B. Andersen.

## Abstract

*Helicobacter pylori* colonizes the human stomach and may affect the inflammatory response, hormone production related to energy regulation, and gut microbiota composition. Previous studies have demonstrated an inverse correlation between *H. pylori* seropositivity and pediatric obesity. We hypothesized that we would find a similar relationship among Danish children and adolescents. We assessed *H. pylori* seroprevalence in 713 subjects from an obesity clinic cohort and 990 subjects from a population-based cohort, and its association with obesity and other cardiometabolic risk factors. No association was found between *H. pylori* and body mass index (BMI) standard deviation score (SDS). *H. pylori* seropositivity was, however, associated with higher fasting blood glucose levels and the prevalence of hyperglycemia, suggesting that *H. pylori* may contribute to impaired glucose regulation in Danish children and adolescents.

**Graphical abstract:** 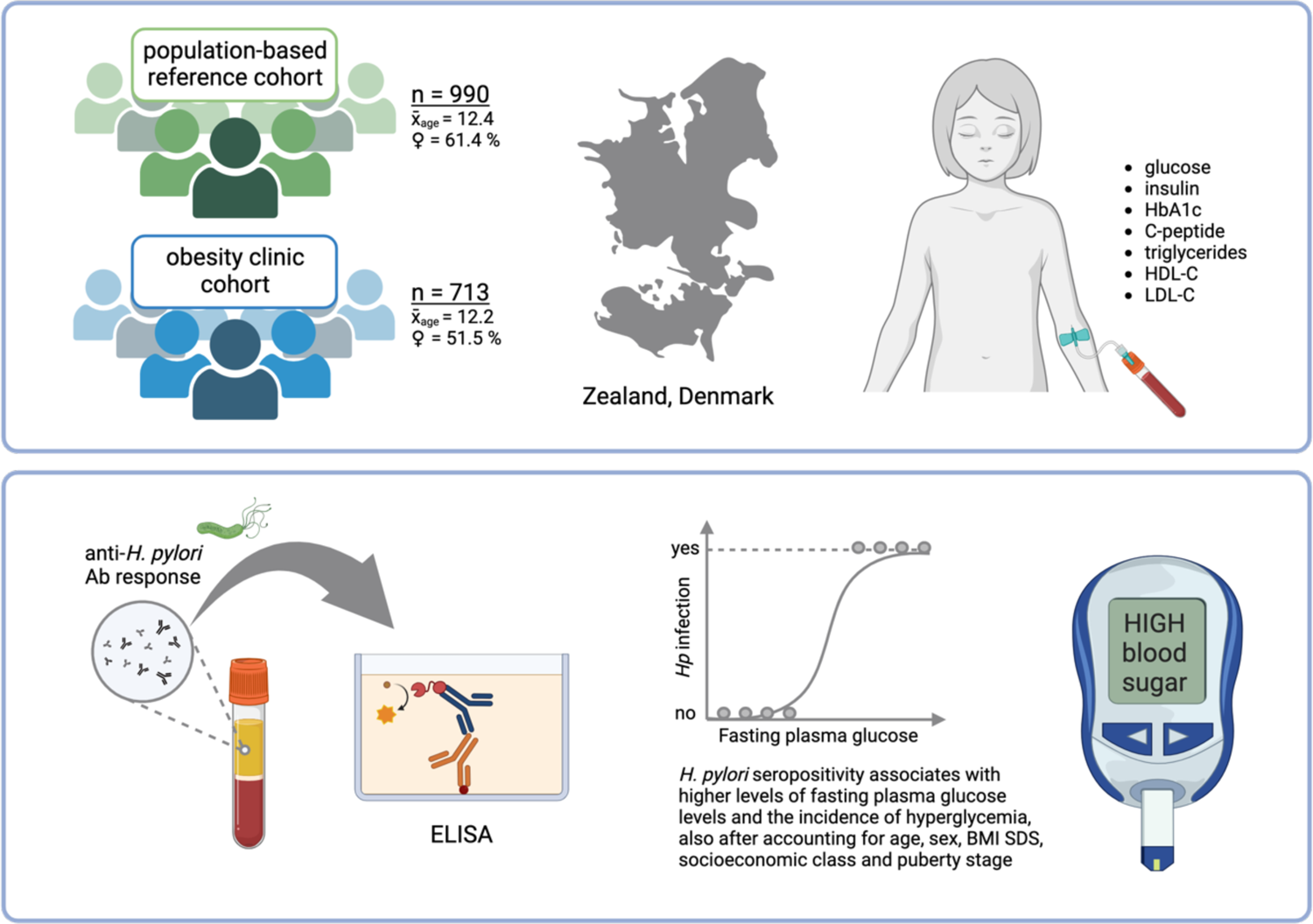

## INTRODUCTION

Obesity is a complex disease defined by excessive or abnormal fat accumulation that may impair health (1). Further, obesity is characterized by chronic low-grade inflammation, which increases the risk of developing cardiometabolic complications (2). The prevalence of obesity has reached epidemic proportions globally and continues to rise. Recently, the World Health Organization (WHO) reported that among children and adolescents, the global prevalence has increased from 4% in 1975 to 18% in 2015 (1). In Denmark, a surge in obesity has been reported among school children in Copenhagen from 1947 to 2003 (3), and nationwide from 1995 to 2002 (4). A follow up study on Copenhagen school children from 2002 to 2007 found a stagnation in the obesity rates among children (from 17.8% to 15.9% in girls, and from 14.0% to 11.6% in boys), but a continuous increase among adolescents (up to 25.4% in girls and 18.9% in boys) (5).

The development of obesity is affected by a complex interplay between genetic, socioeconomic, and environmental factors. Emerging research suggests that the composition of the gastrointestinal microbiome also plays a role in obesity (6,7). Disruption of the early life microbiome, such as a Caesarean section or antibiotic treatment, increase the risk of obesity later in life (8–10). This could be the result of a less diverse microbiome potentially hindering the proper development of the host immune system and triggering the development of autoimmune diseases and obesity (11).

*Helicobacter pylori* is an ancestral member of the gastric microbiome that has been associated with humans for at least 300,000 years (12). While prevalence is declining in association with improving standards of living, *H. pylori* is estimated to colonize about half of the world’s population and is usually acquired during early childhood (13). For most carriers, colonization remains asymptomatic, but persists throughout the host’s lifetime if not treated. Around 20% of infected individuals will develop gastroduodenal malignancies, which primarily manifest in adulthood (14). It is currently not well understood what determines the outcome of a *H. pylori* infection, but age at acquisition (15), presence of virulence factors (16) and host-microbe genomic match (17) seem to play important roles. Mechanistic studies in mice have revealed that *H. pylori* has immunomodulatory effects that can confer protection against asthma and allergies, most robust when the infection is established early in life (18,19). Similar results were found in human correlative studies (20,21).

*H. pylori* has been investigated in relation to pediatric obesity, with two studies showing lower *H. pylori* seropositivity in children and adolescents with obesity (22,23). Other studies found no association (24,25). One European study demonstrated an increase in body mass index (BMI) after eradication of *H. pylori* (26), similar to an earlier finding in adults (27). Results from studies on adults show conflicting results concerning an effect of *H. pylori* on obesity (28). This may be attributed to confounding variables, as both obesity and *H. pylori* infection status is known to correlate with ethnicity, socioeconomic status, age and sex (29–31).

In the present study, we aimed to investigate the relationship between *H. pylori* seropositivity and obesity in Danish children and adolescents and its association with cardiometabolic risk factors. Based on *H. pylori*’s immunomodulatory properties, along with previous findings from pediatric cohorts (22,23,32,33), we hypothesized that *H. pylori* seropositivity will be inversely associated with obesity and cardiometabolic risk factors.

## RESULTS

### *H. pylori* seroprevalence and characteristics of the participants stratified by *H. pylori* seropositivity status

Out of the 1703 included subjects, we found 253 individuals with *H. pylori* seropositivity by whole-cell enzyme-linked immunosorbent assay (ELISA), resulting in a prevalence of 14.8%. Individuals *H. pylori* with seropositivity were older than those with seronegativity (Table 1, *p* = 0.04) and there was a higher proportion of those with seropositivity than seronegativity in the lowest socioeconomic status (*p* = 0.015). Subjects with *H. pylori* seropositivity constituted a larger proportion in the hyperglycemic group (*p* < 0.001) and had higher fasting plasma glucose (*p* = 0.006), serum insulin (*p* = 0.018) and homeostatic model assessment for insulin resistance (HOMA-IR) concentrations (*p* = 0.007) than subjects expressing seronegativity. When analysing the two cohorts separately, we observed that the association between *H. pylori* seropositivity and socioeconomic status was only statistically significant within the obesity clinic cohort (Table S1). Meanwhile, the association between *H. pylori* seropositivity and hyperglycemia, as well as the correlations with fasting plasma glucose and HOMA-IR, were only significant in the population-based reference cohort (Table S2). Within the population-based reference cohort, we found a significant association between *H. pylori* seropositivity and passive smoking (Table S2). Overall, 71 individuals were identified with genotypes other than European of which 26.7 % exhibited *H. pylori* seropositivity and 81.7 % had obesity. Excluding the 71 subjects with non-European ethnicities from the dataset resulted in a weaker and non-significant association between *H. pylori* seropositivity and age, as well as socioeconomic status (Table S3). Furthermore, after exclusion of an additional 159 subjects who self-reported their ethnicity as non-Danish, we found a significant association solely between *H. pylori* seropositivity and fasting plasma glucose levels (Table S4).

**Table 1:**
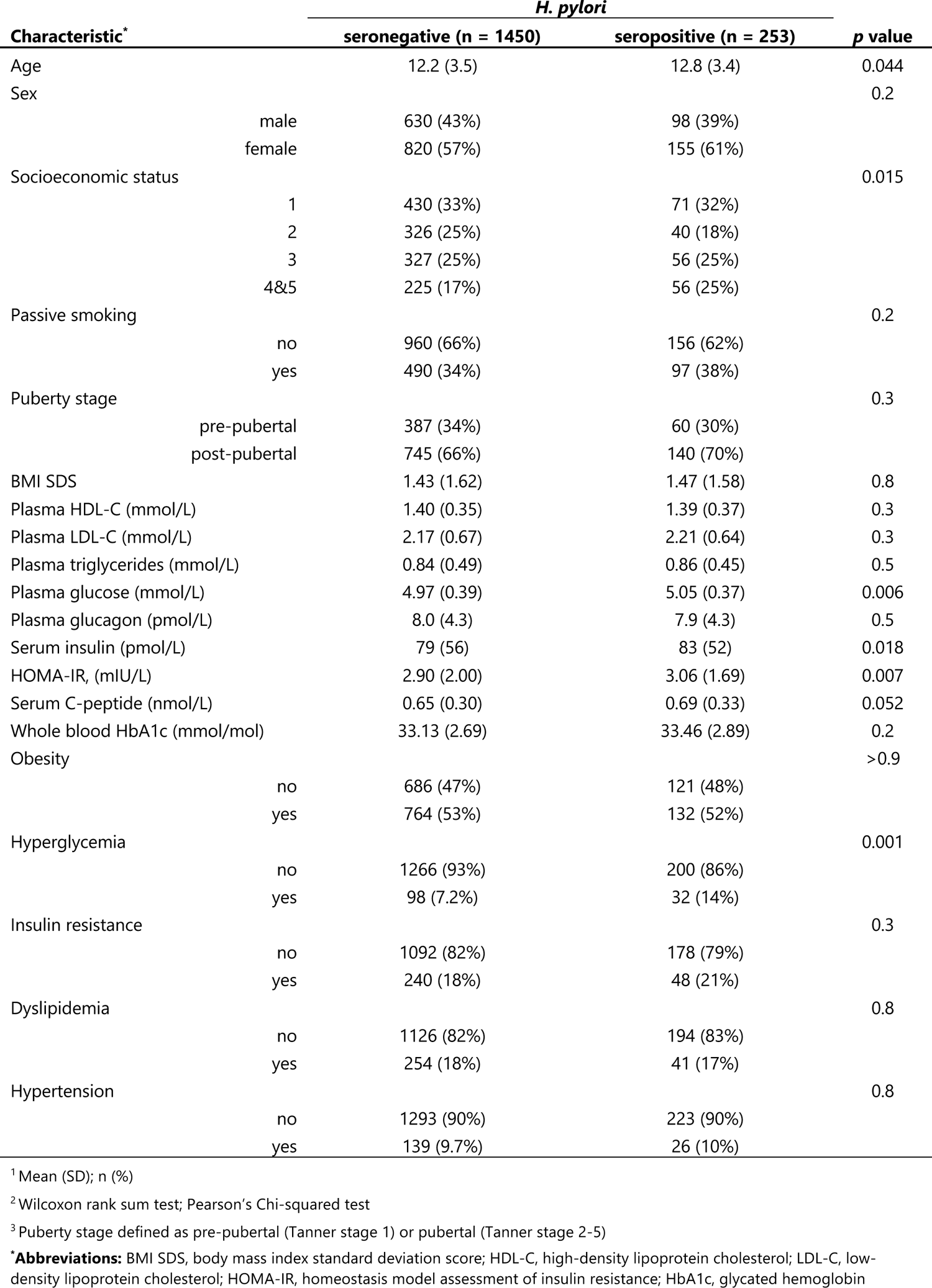
Descriptive characteristics of study population stratified by *Helicobacter pylori* infection status.

As children physiologically have lower antibody titers than adults, we attempted to adjust our adult-based cut-off value by validating a subset (n = 60) of our samples with multiplex serology. The seropositivity rate was overall lower in multiplex serology, compared to ELISA. However, all eight samples identified as seropositive by multiplex serology were also classified as positive or intermediate in the ELISA assay (Table S5). Since multiplex serology was less sensitive in the detection of *H. pylori* antibodies in children’s and adolescents’ sera compared to ELISA, we decided to base the analyses in this study on the adult-based validated ELISA cut-off value.

### *H. pylori* infection as a predictor for cardiometabolic risk factors

*H. pylori* seropositivity was associated with a higher incidence of hyperglycemia (Table 2, model 1, OR = 2.03, *p* = 0.001), when adjusted for age, sex and BMI standard deviation score (SDS), and also after additional adjustment for socioeconomic status (model 2, OR = 1.96, *p* = 0.005) and puberty stage (model 3, OR = 2.36, *p* < 0.001). Among the cardiometabolic risk features tested, *H. pylori* seropositivity was associated with higher levels of fasting plasma glucose (Table 3, model 1, β = 0.2, *p* = 0.005) when adjusted for age, sex and BMI SDS, but also after additional adjustment of socioeconomic status (model 2, β = 0.16, *p* = 0.03) and puberty stage (model 3, β = 0.2, *p* = 0.016).

**Table 2:** Estimated odds ratios (ORs) with 95 % confidence intervals (CI) for associations of *Helicobacter pylori* seropositivity as an indicator of categorical (yes/no) cardiometabolic risk factors.

|  | Model 1<sup>a</sup> |  |  | Model 2<sup>b</sup> |  |  | Model 3<sup>c</sup> |  |  |
| --- | --- | --- | --- | --- | --- | --- | --- | --- | --- |
|  | n* | OR (95% CI) | p | n* | OR (95% CI) | p | n* | OR (95% CI) | p |
| Obesity | 1703 | 1.01 (0.77;1.32) | 0.94 | 1531 | 0.85 (0.62;1.17) | 0.32 | 1209 | 0.94 (0.65;1.35) | 0.73 |
| Hyperglycemia | 1596 | 2.03 (1.30;3.09) | 0.001 | 1436 | 1.96 (1.20;3.10) | 0.005 | 1151 | 2.36 (1.40;3.88) | 0.001 |
| Hypertension | 1681 | 1.08 (0.67;1.69) | 0.73 | 1514 | 0.86 (0.50;1.43) | 0.58 | 1199 | 0.96 (0.54;1.64) | 0.90 |
| Dyslipidemia | 1615 | 0.87 (0.58;1.29) | 0.50 | 1455 | 0.88 (0.57;1.35) | 0.57 | 1164 | 0.79 (0.47;1.27) | 0.34 |
| Insulin resistance | 1558 | 1.26 (0.85;1.84) | 0.24 | 1404 | 1.13 (0.73;1.72) | 0.57 | 1122 | 1.36 (0.84;2.17) | 0.20 |

<sup>a</sup>adjusted for age, sex, BMI SDS (except for the outcome "obesity")
<sup>b</sup>additional adjustment for socioeconomic status
<sup>c</sup>additional adjustment for puberty stage, defined as pre-pubertal (Tanner stage 1) or pubertal (Tanner stage 2-5)
\*sample size after removal of missing values

**Table 3:** Standardized coefficient (β) estimates with 95 % confidence intervals (CI) for associations of *Helicobacter pylori* seropositivity as an indicator of continuous cardiometabolic risk factors. Outcome variables were log10-transformed and z-scored, except for BMI standardized deviation score (SDS), bodyfat % SDS and waist-to-height ratio (WtHR).

|  | Model 1<sup>a</sup> |  |  | Model 2<sup>b</sup> |  |  | Model 3<sup>c</sup> |  |  |
| --- | --- | --- | --- | --- | --- | --- | --- | --- | --- |
| | n* | $\beta$ (95% CI) | p | n* | $\beta$ (95% CI) | p | n* | $\beta$ (95% CI) | p |
| BMI SDS | 1703 | 0.07 (-0.14;0.29) | 0.50 | 1531 | -0.04 (-0.24;0.17) | 0.73 | 1209 | 0.01 (-0.21;0.23) | 0.92 |
| Body fat % SDS | 698 | -0.02 (-0.20;0.16) | 0.82 | 609 | -0.01 (-0.19;0.18) | 0.94 | 474 | -0.05 (-0.26;0.15) | 0.61 |
| WtHR SDS | 1610 | 0.013 (-0.17;0.19) | 0.89 | 1452 | -0.077 (-0.25;0.10) | 0.38 | 1155 | -0.05 (-0.24;0.13) | 0.57 |
| Glucose | 1602 | 0.20 (0.06;0.33) | 0.005 | 1441 | 0.16 (0.01;0.31) | 0.03 | 1156 | 0.20 (0.04;0.36) | 0.016 |
| Insulin | 1626 | 0.08 (-0.03;0.19) | 0.18 | 1462 | 0.05 (-0.06;0.17) | 0.38 | 1171 | 0.09 (-0.04;0.22) | 0.17 |
| HOMA-IR | 1595 | 0.11 (-0.01;0.22) | 0.07 | 1434 | 0.07 (-0.05;0.20) | 0.25 | 1150 | 0.11 (-0.03;0.24) | 0.11 |
| HbA1c | 1613 | 0.005 (-0.0004;0.01) | 0.07 | 1453 | 0.003 (-0.002;0.01) | 0.25 | 1164 | 0.06 (-0.10;0.22) | 0.43 |
| C-peptide | 1582 | 0.06 (-0.04;0.17) | 0.25 | 1164 | 0.06 (-0.05;0.17) | 0.28 | 1150 | 0.10 (-0.02;0.22) | 0.09 |

<sup>a</sup>adjusted for age, sex, BMI SDS (except for the outcome "BMI SDS", "Body fat % SDS" and "WtHR SDS")
<sup>b</sup>additional adjustment for socioeconomic status
<sup>c</sup>additional adjustment for puberty stage, defined as pre-pubertal (Tanner stage 1) or pubertal (Tanner stage 2-5)
\*sample size after removal of missing values

As a sensitivity analysis, we lowered the cut-off value for *H. pylori* seropositivity to see how it affected the model outputs. The cut-off value could be lowered to 0.7 without changing the logistic and linear regression model outputs markedly (Table S6 and S7). For every ten percent we lowered the *H. pylori* seropositivity cut-off value, the seroprevalence increased about 2% (Table S6). Excluding subjects with genetic ethnicities other than European resulted in stronger associations between *H. pylori* seropositivity and fasting plasma glucose levels, as well as the occurrence of hyperglycemia (Table S8 and S9). Additional exclusion of subjects with self-reported ethnicity other than Danish led to weaker associations between *H. pylori* seropositivity and both fasting plasma glucose levels and the prevalence of hyperglycemia (Table S10 and S11).

## DISCUSSION

Opposed to our hypothesis, we did not find an association between *H. pylori* seropositivity and pediatric obesity. BMI does have several shortcomings as a proxy for obesity, such as the inability to differentiate between fat, bone, and muscle or the changes in body fat composition depending on age and sex (34). Dual-energy x-ray absorptiometry (DEXA) offers a solution to the first issue mentioned, but this was mainly performed on the obesity clinic cohort, and only on few individuals from the population-based reference cohort. For the individuals where DEXA was performed, we could not detect an association between *H. pylori* seropositivity and body fat % SDS. Waist circumference was available for nearly all study subjects and waist-to-height ratio (WtHR) has been shown be a better indicator of abdominal obesity and metabolic disease than BMI in children (35,36). However, neither for WtHR SDS did we find an association with *H. pylori* infection status.

Interestingly, *H. pylori* seropositivity was associated with higher prevalence of hyperglycemia and higher concentrations of fasting plasma glucose (Table 2 and 3). Hyperglycemia is a condition defined as excessive amounts of circulating blood glucose and mainly occurs in people with diabetes. Of note, no individuals diagnosed with diabetes were identified in these subsets of the HOLBAEK study and those with blood glucose and/or HbA1c values in the diabetic range were excluded. *H. pylori* has been shown in a meta-analysis to be a risk factor for type 2 diabetes (37), which most commonly develops in adulthood. However, the increasing obesity rates among children has led to more pediatric type 2 diabetes cases. Additionally, several studies have found a higher prevalence of *H. pylori* among children with type 1 diabetes (38–40). Another risk factor for diabetes is higher circulating insulin levels, but we did not find any association between *H. pylori* and insulin or pro-insulin (C-peptide) concentrations. Concentrations of HbA1c are commonly used to monitor diabetes treatment but can also be useful in evaluating prediabetes. A large cross-sectional study reported a higher mean level of HbA1c in adults with seropositivity for *H. pylori*, also in a synergistic effect between *H. pylori* and BMI (41). We found a tendency towards higher HbA1c concentrations in subjects with seropositivity for *H. pylori* after adjusting for sex, age and BMI SDS, but this association disappeared after additional adjustment for socioeconomic status and puberty stage (Table 3).

With our somewhat conservative detection threshold we found an *H. pylori* prevalence of 14.8% in the study population, with a significantly higher age of individuals with *H. pylori* seropositivity. No recent surveys on *H. pylori* seropositivity in Denmark have been published, but an epidemiologic study conducted from 1982 to 1984 including 4581 adults aged 30-60 years found an overall *H. pylori* prevalence of 25% (42). In 1998-1999 a population-based *H. pylori* screening disclosed a prevalence of 17% in adults (43). Hence, our results are in line with an expected overall decrease in prevalence (44). Several studies have reported low sensitivities when using ELISAs developed for diagnosis of *H. pylori* infection in adults on children (45,46). We attempted to adjust our adult-based cut-off value with multiplex serology, but compared to ELISA, the sensitivity was even lower. A possible explanation for this discrepancy could be the use of whole-cell derived antigens in the ELISA, as opposed to the multiplex serology that specifically targets antibodies against 12 selected *H. pylori* antigens. These 12 antigens may be less involved in the early immune response to *H. pylori* in children and adolescents compared to adults.

In human studies it is difficult to completely control for confounding variables. Our results confirmed that *H. pylori* seropositivity is associated with lower socioeconomic status, as previously reported (47). This association may be related to ethnicity, as immigrants have a lower socioeconomic status in Denmark (48) and those from non-Western countries would be expected to have a higher *H. pylori* prevalence. Indeed, when we excluded individuals with non-European and non-Danish ethnicities, the association between socioeconomic status and *H. pylori* seropositivity lost significance. We could not detect a significant correlation with sex, although a higher proportion of girls exhibited seropositivity in this study. Previously, a meta-analysis concluded that there is a global male predominance of *H. pylori* infection in adults, but not in children. The authors speculated if this discrepancy is due to differential antibiotic exposure among sexes (49). A later conducted meta-analysis disclosed a small effect of male sex also in children, but the mechanism by which sex influences *H. pylori* infection remains unknown (50).

Our study exhibits notable strengths and limitations. In contrast to prior reports, the surveyed cohorts cover a large number of individuals with comprehensive sociodemographic and cardiometabolic profiles. Additionally, the study participants were recruited for an obesity study, rather than being selected based on gastroduodenal disease symptoms often related to *H. pylori* infection. One limitation of our study is the absence of data related to birth delivery methods (vaginal or C-section), as well as information concerning breast versus formula feeding and antibiotic usage. These factors are recognized contributors to obesity and metabolic disease (51–53). Additionally, we found a low prevalence of *H. pylori* at 14.8%, which may make it more challenging to detect associations compared to in high prevalence populations. A large proportion of our study population had been genotyped, facilitating the systematic removal of individuals with non-European ethnicity to assess its impact on our analysis. Nonetheless, we acknowledge the possibility that among the subset of subjects with missing genotypes, there may be individuals with non-European ethnicity. Out of the 324 subjects with missing genotypes, 280 had self-reported their ethnicity, and we therefore ran the analyses with additional exclusion of individuals with non-Danish ethnicity. However, this criterion is less objective and may exclude subjects with European ethnicity.

In conclusion, we find no association between *H. pylori* seropositivity and obesity in Danish children and adolescents. *H. pylori* seropositivity was associated with the incidence of hyperglycemia and higher fasting plasma glucose concentrations. In support of our findings, a recent experimental study revealed that *H. pylori* worsens the impaired glucose regulation in mice on a high fat diet (54). Further, our own experimental work on mice fed a high fat diet showed that early-life *H. pylori* infection worsens the metabolic state and perturbs the distal gut microbiome composition (unpublished results). Taken together, we suggest that the presence of *H. pylori* in children and adolescents can impair glucose metabolism. As such, loss of this ancient microbial associate may be beneficial for early life host metabolism in the Scandinavian population and environment.

## METHODS

### Study populations

We used a random subset of individuals from The HOLBAEK Study, that has been described elsewhere (55,56). Briefly, serum samples derived from two groups of children and adolescents: (a) an obesity clinic group (n = 713) consisting of individuals with a BMI SDS > 90th percentile (BMI SDS > 1.28) according to Danish reference values (57), who followed a multifaceted childhood obesity management program at Holbæk Hospital and (b) a population-based reference group (n = 990) consisting of subjects recruited from schools in the same geographical area. Participants from both groups were enrolled in the HOLBAEK study between 2010 and 2019. The study participants have been thoroughly characterized in relation to their age, sex, socioeconomic status and various cardiometabolic risk factors. The characteristics of the full obesity clinic and population-based cohorts have been described elsewhere (56). In brief, the study groups did not differ in age, but there were more boys in the obesity-clinic cohort (45.8%) than in the population-based cohort (40.3%). In the present study, exclusion criteria were children under six (n = 10) and over 19 years of age (n = 13). Additionally, subjects who fulfilled type 2 diabetes criteria (58) based on blood samples showing fasting plasma glucose > 7.0 mmol/L (n = 2) and/or glycated hemoglobin (HbA1c) > 48 mmol/mol (n = 2) were excluded.

### Ethics

In accordance with the Declaration of Helsinki, participants provided informed consent. For individuals below 18 years of age, written consent was acquired from their parents or legal guardians, whereas those 18 years or older provided their own written consent. The study received approval from the Scientific Ethics Committee of Region Zealand, Denmark (protocol No. SJ-104) and the Danish Data Protection Agency. The HOLBAEK Study is registered at ClinicalTrials.gov (NCT00928473).

### Anthropometrics

Anthropometric data (height, weight, and waist measurements) was collected during routine clinical examinations in the obesity clinic group. Meanwhile, the population-based group underwent assessments in a mobile laboratory, administered by trained medical professionals. WtHR SDS was calculated based on age- and sex-specific reference values (59). Whole-body dual-energy x-ray absorptiometry (DEXA) was performed to quantify body fat % in a subset from both the obesity clinic (n = 650) and population-based (n = 71) groups, using a GE Lunar iDXA (ME+200179, GE Healthcare) as earlier described (60). Body fat % SDS was calculated based on age- and sex-specific reference values (61). Puberty stage was assessed by the Tanner stage classification method (62,63). In the obesity clinic group, a pediatrician evaluated Tanner stages based on breast development in girls and gonad development in boys. In the population-based group, a self-assessment approach was utilized, where individuals used a standard questionnaire with picture patterns for recognition.

### Genotyping

Participant genotyping was carried out as described previously (64,65). Out of the 1703 included participants, 1379 had been genotyped. In brief, DNA was extracted and genotyped by using Illumina Infinium HumanCoreExome-12 v1.0 and HumanCoreExome-24 v1.1 Beadchips, with the Illumina HiScan system. Genotype calling was performed using the Genotyping module (version 1.9.4) of GenomeStudio software (version 2011.1; Illumina). Subsequently, the data were phased using EAGLE2 (version 2.0.5) and imputed to the Haplotype Reference Consortium (HRC, r1.1) using PBWT on the Sanger server. Individuals not of European descent were identified through Principal Component Analysis (PCA) using ancestry informative markers. Study samples were classified as non-European when their Euclidean distance from the center exceeded a radius greater than 1.5 times the maximum Euclidean distance of the European reference samples from the 1000 Genomes dataset (66).

### Biochemical analyses

Serum was sampled as previously described (67). Briefly, venous blood samples were collected from 7 to 9 am in ice-cold EDTA tubes following an overnight fast of minimum 8 hours and separated by centrifugation within 20 minutes. Samples were stored at −80 °C until further analysis. Serum antibody responses to *H. pylori* were determined by ELISA as previously described (68) with a few modifications. *H. pylori* whole-cell antigen was collected from the *H. pylori* strain PMSS1 (69) grown on blood agar plates for 48h under microaerophilic conditions and lyzed with the B-PER™ Bacterial Protein Extraction Kit (Fisher Scientific). Antigens were coated to microtiter wells (Immulon 2 HB) in carbonate-bicarbonate buffer (0.05M, pH 9.6) to a final concentration of 0.1 mg/well at room temperature (RT) overnight. The following morning, plates were blocked for 3h at 37°C with 200 uL phosphate-buffered saline (PBS, pH 7.2-7.4) containing 0.5% Tween 20 and 0.1% gelatin (PBSTG). Subsequently, plates were washed twice with 250 uL PBS containing 0.5% Tween 20 (PBST). Serum samples were diluted 1:200 in 100 uL PBSTG containing 0.5% bovine γ-globulin (PBSTGG) and analyzed in duplicate. On each plate, four positive serum controls and four negative serum controls were included, along with two blanks and four calibrator samples from the *H. pylori* IgG Test System (Zeus Scientific). The positive serum control consisted of a pool of 30 samples from the HOLBAEK study that were determined as seropositive with the *H. pylori* IgG Test System (Zeus Scientific). After 1h incubation at 37°C, plates were washed three times with 250 uL PBST per well. Next, peroxidase labelled antibody (polyclonal IgG Fc Cross-Adsorbed Goat anti-Human, Invitrogen) was diluted 1:500 in 100 uL PBST containing 0.1% bovine γ-globulin and 1% bovine serum albumin (PBSTGB) and incubated for 1h at 37°C. Finally, plates were washed five times with 250 uL PBST and then incubated with 100 uL substrate solution (Mcllvains buffer pH 4.6 with 0.1% ABTS and 0.16% H_2_O_2_ [30%]) for 5 min at RT in the dark before absorbance was measured at 420 nm. Optical density (OD) ratio was calculated by standardizing the absorbance values with the mean calibrator value. Pre-defined cut-off values were as follows: OD ratio < 0.2, negative; 0.2 ≤ OD ratio < 1, intermediate; OD ratio ≥ 1, positive.

A subset of 60 samples (n = 6 negatives, n = 44 intermediate, and n = 10 positive samples) was re-analyzed by multiplex serology to validate the adult-based pre-defined cut-off in children and adolescent’s sera with an independent assay (45,70). It was ensured that cohort, age and sex distribution resembled the total study population. *H. pylori* multiplex serology was performed as previously described (71). Briefly, 12 *H. pylori* proteins were recombinantly expressed as glutathione-S-transferase (GST)-tagged proteins and affinity-purified on glutathione-casein coated beads with distinct internal fluorescence (Luminex Corp., Austin, TX, USA). Sera were incubated with a suspension array of the antigen-loaded beads in a 1:100 serum dilution and bound serum IgG serum antibodies were labelled using a biotinylated goat anti-human IgG secondary antibody (Jackson ImmunoResearch, Ely, UK) and Streptavidin-R-phycoerythrin (Moss Inc., Pasadena, MD, USA). A Luminex analyzer (Luminex Corp., Austin, TX, USA) then identifies the bead type and consequently the bound antigen as well as quantifies the amount of bound serum antibodies given as the median fluorescence intensity of 100 beads per type measured. Pre-defined antigen-specific cut-offs for *H. pylori* antigens were applied (Table S12) and quality assured by the visual inflection point method, and samples were classified as overall *H. pylori* seropositive when being positive to any 4 or more of the included *H. pylori* antigens as described previously (72).

### Definition of variables

For the purpose of this study, obesity was defined as a BMI SDS ≥ 90^th^ percentile. Definition of the remaining cardiometabolic risk factors (hyperglycemia, hypertension, dyslipidemia and insulin resistance) has been described elsewhere (56). Age was included as a continuous variable. Tanner stage was dichotomized, where Tanner stage 1 was classified as pre-pubertal and Tanner 2-5 was classified as pubertal/post-pubertal. The socioeconomic status variable was categorized into five levels according to the working status of the parents or legal guardians, with 1 representing highest level (general directors and others), 2 higher middle level (managers, company owners and others), 3 middle level (functionaries, skilled workers and others), 4 lower middle level (workers, students and others) and 5 lowest level (unemployed). In this study, socioeconomic status level 4 and 5 were merged to increase statistical power because of small sample sizes. Ethnicity as self-reported country of origin and ancestry (available for 1558 out of 1703 participants) was categorized as Danish or non-Danish.

### Statistical analyses

All statistical analyses were performed in R studio (version 4.1.1). Continuous variables were characterized as means with standard deviation (SD) and categorical variables were described as frequencies and percentages (%). *P* values for continuous variables comparing two groups were calculated using Wilcoxon rank sum tests, while categorical variables were calculated using chi-square tests. Generalized logistic regression models were used to calculate odds ratios (OR) and 95% confidence intervals (CI) with *H. pylori* infection status (0/1) as predictor of cardiometabolic risk factors (obesity, hypertension, hyperglycemia, insulin resistance and dyslipidemia) with the adjustment of age, sex, BMI SDS (except when the outcome was obesity). A second model was calculated with the additional adjustment of socioeconomic status, and a third model was additionally adjusted for puberty stage to account for transient insulin resistance (73). Linear regression models were used to calculate the effect size (β) and 95% confidence intervals (CI) with *H. pylori* infection status (0/1) as predictor of BMI SDS, body fat % SDS, WtHR SDS, fasting plasma glucose, fasting serum insulin, fasting whole blood HbA1c and fasting serum C-peptide (all variables were log10 transformed and z-scored prior to analysis, except BMI SDS, body fat % SDS and WtHR SDS) with the adjustment of age, sex and BMI SDS (except when the outcome was BMI SDS, body fat % SDS or WtHR SDS). A second model was additionally adjusted for socioeconomic status and a third model for puberty stage. Missing values were removed prior to the analysis.

## Data Availability

Restrictions apply to the availability of some or all data generated or analyzed during this study to preserve patient confidentiality. The corresponding author will on request detail the restrictions and any conditions under which access to some data may be provided.

