## Supplementary tables for "*Helicobacter pylori* seropositivity associates with hyperglycemia, but not obesity in Danish children and adolescents"

**Table S1:** Descriptive characteristics of obesity clinic cohort stratified by *Helicobacter pylori* infection status

| **Characteristic*** | | ***H. pylori*[1]** | | |
|---|---|---|---|---|
| | | **seronegative (n = 600)** | **seropositive (n = 113)** | ***p* value[2]** |
| Age | | 12.1 (3.1) | 12.5 (3.2) | 0.4 |
| Sex | | | | 0.3 |
| | male | 297 (50%) | 49 (43%) | |
| | female | 303 (50%) | 64 (57%) | |
| Socioeconomic status | | | | 0.029 |
| | 1 | 46 (9.0%) | 12 (13%) | |
| | 2 | 133 (26%) | 13 (14%) | |
| | 3 | 166 (32%) | 28 (30%) | |
| | 4&5 | 167 (33%) | 41 (44%) | |
| Passive smoking | | | | 0.6 |
| | no | 287 (48%) | 58 (51%) | |
| | yes | 313 (52%) | 55 (49%) | |
| Puberty stage[3] | | | | >0.9 |
| | pre-pubertal | 185 (41%) | 37 (42%) | |
| | post-pubertal | 262 (59%) | 52 (58%) | |
| BMI SDS | | 3.00 (0.69) | 2.95 (0.71) | 0.4 |
| Plasma HDL-C (mmol/L) | | 1.23 (0.31) | 1.20 (0.30) | 0.2 |
| Plasma LDL-C (mmol/L) | | 2.37 (0.70) | 2.39 (0.59) | 0.4 |
| Plasma triglycerides (mmol/L) | | 1.08 (0.59) | 1.10 (0.49) | 0.4 |
| Plasma glucose (mmol/L) | | 4.94 (0.40) | 4.98 (0.36) | 0.11 |
| Plasma glucagon (pmol/L) | | 9.9 (4.6) | 9.4 (5.1) | 0.2 |
| Serum insulin (pmol/L) | | 102 (73) | 104 (66) | 0.2 |
| HOMA-IR, (mIU/L) | | 3.67 (2.54) | 3.67 (1.98) | 0.3 |
| Serum C-peptide (nmol/L) | | 0.77 (0.39) | 0.81 (0.44) | 0.3 |
| Whole blood HbA1c (mmol/mol) | | 33.3 (2.9) | 33.9 (3.1) | 0.2 |
| Obesity | | | | >0.9 |
| | no | 1 (0.2%) | 0 (0%) | |
| | yes | 599 (100%) | 113 (100%) | |
| Hyperglycemia | | | | 0.5 |
| | no | 483 (91%) | 87 (88%) | |
| | yes | 50 (9.4%) | 12 (12%) | |
| Insulin resistance | | | | 0.6 |
| | no | 361 (69%) | 63 (66%) | |
| | yes | 165 (31%) | 33 (34%) | |
| Dyslipidemia | | | | 0.5 |
| | no | 354 (66%) | 69 (70%) | |
| | yes | 184 (34%) | 30 (30%) | |
| Hypertension | | | | >0.9 |
| | no | 477 (82%) | 90 (83%) | |
| | yes | 105 (18%) | 19 (17%) | |

[1]Mean (SD); n (%)

[2]Wilcoxon rank sum test; Pearson's Chi-squared test

[3]Puberty stage defined as pre-pubertal (Tanner stage 1) or pubertal (Tanner stage 2-5)

***Abbrevations:** BMI SDS, body mass index standard deviation score; HDL-C, high-density lipoprotein cholesterol; LDL-C, low-density lipoprotein cholesterol; HOMA-IR, homeostasis model assessment of insulin resistance; HbA1c, glycated hemoglobin

**Table S2**: Descriptive characteristics of population-based reference cohort stratified by *Helicobacter pylori* infection status

| Characteristic* | | *H. pylori*[1] | | *p* value[2] |
|---|---|---|---|---|
| | | seronegative (n = 850) | seropositive (n = 140) | |
| Age | | 12.3 (3.7) | 13.0 (3.5) | 0.067 |
| Sex | | | | 0.4 |
| | male | 333 (39%) | 49 (35%) | |
| | female | 517 (61%) | 91 (65%) | |
| Socioeconomic status | | | | 0.3 |
| | 1 | 384 (48%) | 59 (46%) | |
| | 2 | 193 (24%) | 27 (21%) | |
| | 3 | 161 (20%) | 28 (22%) | |
| | 4&5 | 58 (7.3%) | 15 (12%) | |
| Passive smoking | | | | 0.021 |
| | no | 673 (79%) | 98 (70%) | |
| | yes | 177 (21%) | 42 (30%) | |
| Puberty stage[3] | | | | 0.074 |
| | pre-pubertal | 202 (29%) | 23 (21%) | |
| | post-pubertal | 483 (71%) | 88 (79%) | |
| BMI SDS | | 0.33 (1.08) | 0.28 (0.94) | 0.7 |
| Plasma HDL-C (mmol/L) | | 1.52 (0.33) | 1.53 (0.35) | 0.8 |
| Plasma LDL-C (mmol/L) | | 2.04 (0.62) | 2.08 (0.64) | 0.5 |
| Plasma triglycerides (mmol/L) | | 0.69 (0.32) | 0.68 (0.33) | 0.8 |
| Plasma glucose (mmol/L) | | 4.99 (0.38) | 5.09 (0.37) | 0.014 |
| Plasma glucagon (pmol/L) | | 6.8 (3.6) | 6.7 (3.2) | 0.9 |
| Serum insulin (pmol/L) | | 64 (33) | 67 (30) | 0.076 |
| HOMA-IR, (mIU/L) | | 2.41 (1.35) | 2.61 (1.26) | 0.034 |
| Serum C-peptide (nmol/L) | | 0.58 (0.21) | 0.61 (0.20) | 0.2 |
| Whole blood HbA1c (mmol/mol) | | 33.04 (2.56) | 33.17 (2.68) | 0.8 |
| Obesity | | | | 0.13 |
| | no | 685 (81%) | 121 (86%) | |
| | yes | 165 (19%) | 19 (14%) | |
| Hyperglycemia | | | | <0.001 |
| | no | 783 (94%) | 113 (85%) | |
| | yes | 48 (5.8%) | 20 (15%) | |
| Insulin resistance | | | | 0.5 |
| | no | 731 (91%) | 115 (88%) | |
| | yes | 75 (9.3%) | 15 (12%) | |
| Dyslipidemia | | | | >0.9 |
| | no | 772 (92%) | 125 (92%) | |
| | yes | 70 (8.3%) | 11 (8.1%) | |
| Hypertension | | | | 0.7 |
| | no | 816 (96%) | 133 (95%) | |
| | yes | 34 (4.0%) | 7 (5.0%) | |

[1]Mean (SD); n (%)

[2]Wilcoxon rank sum test; Pearson's Chi-squared test

[3]Puberty stage defined as pre-pubertal (Tanner stage 1) or pubertal (Tanner stage 2-5)

***Abbrevations:** BMI SDS, body mass index standard deviation score; HDL-C, high-density lipoprotein cholesterol; LDL-C, low-density lipoprotein cholesterol; HOMA-IR, homeostasis model assessment of insulin resistance; HbA1c, glycated hemoglobin

**Table S3:** Descriptive characteristics of study population stratified by *Helicobacter pylori* infection status. 71 subjects with non-European ethnicity were excluded.

| | | ***H. pylori***[1] | | |
|---|---|---|---|---|
| **Characteristic*** | | **seronegative (n = 1398)** | **seropositive (n = 234)** | ***p* value**[2] |
| Age | | 12.3 (3.5) | 12.7 (3.4) | 0.075 |
| Sex | | | | 0.14 |
| | male | 601 (43%) | 88 (38%) | |
| | female | 797 (57%) | 146 (62%) | |
| Socioeconomic status | | | | 0.083 |
| | 1 | 425 (33%) | 70 (33%) | |
| | 2 | 314 (25%) | 39 (18%) | |
| | 3 | 325 (26%) | 56 (26%) | |
| | 4&5 | 208 (16%) | 47 (22%) | |
| Passive smoking | | | | 0.2 |
| | no | 928 (66%) | 144 (62%) | |
| | yes | 470 (34%) | 90 (38%) | |
| Puberty stage[3] | | | | 0.2 |
| | pre-pubertal | 374 (34%) | 56 (30%) | |
| | post-pubertal | 716 (66%) | 133 (70%) | |
| BMI SDS | | 1.40 (1.61) | 1.35 (1.55) | 0.6 |
| Plasma HDL-C (mmol/L) | | 1.40 (0.35) | 1.40 (0.37) | 0.5 |
| Plasma LDL-C (mmol/L) | | 2.17 (0.67) | 2.21 (0.64) | 0.3 |
| Plasma triglycerides (mmol/L) | | 0.84 (0.49) | 0.84 (0.45) | 0.8 |
| Plasma glucose (mmol/L) | | 4.97 (0.39) | 5.05 (0.37) | 0.003 |
| Plasma glucagon (pmol/L) | | 7.9 (4.2) | 7.7 (3.9) | 0.4 |
| Serum insulin (pmol/L) | | 78 (54) | 81 (52) | 0.055 |
| HOMA-IR, (mIU/L) | | 2.87 (2.00) | 2.99 (1.67) | 0.022 |
| Serum C-peptide (nmol/L) | | 0.65 (0.30) | 0.68 (0.34) | 0.2 |
| Whole blood HbA1c (mmol/mol) | | 33.09 (2.66) | 33.38 (2.86) | 0.3 |
| Obesity | | | | 0.5 |
| | no | 675 (48%) | 119 (51%) | |
| | yes | 723 (52%) | 115 (49%) | |
| Hyperglycemia | | | | <0.001 |
| | no | 1,224 (93%) | 184 (86%) | |
| | yes | 91 (6.9%) | 30 (14%) | |
| Insulin resistance | | | | 0.4 |
| | no | 1,062 (83%) | 167 (80%) | |
| | yes | 221 (17%) | 42 (20%) | |
| Dyslipidemia | | | | 0.5 |
| | no | 1,086 (82%) | 182 (84%) | |
| | yes | 244 (18%) | 35 (16%) | |
| Hypertension | | | | 0.8 |
| | no | 1,247 (90%) | 210 (91%) | |
| | yes | 136 (9.8%) | 21 (9.1%) | |

[1]Mean (SD); n (%)

[2]Wilcoxon rank sum test; Pearson's Chi-squared test

[3]Puberty stage defined as pre-pubertal (Tanner stage 1) or pubertal (Tanner stage 2-5)
***Abbrevations:** BMI SDS, body mass index standard deviation score; HDL-C, high-density lipoprotein cholesterol; LDL-C, low-density lipoprotein cholesterol; HOMA-IR, homeostasis model assessment of insulin resistance; HbA1c, glycated hemoglobin

**Table S4:** Descriptive characteristics of study population stratified by *Helicobacter pylori* infection status. 71 subjects with non-European genetic ethnicity and 159 subjects with self-reported non-Danish ethnicity were excluded.

| | | ***H. pylori*[1]** | | |
|---|---|---|---|---|
| **Characteristic*** | | **seronegative (n = 1276)** | **seropositive (n = 197)** | ***p* value[2]** |
| Age | | 12.3 (3.5) | 12.6 (3.4) | 0.4 |
| Sex | | | | 0.077 |
| | male | 549 (43%) | 71 (36%) | |
| | female | 727 (57%) | 126 (64%) | |
| Socioeconomic status | | | | 0.11 |
| | 1 | 404 (35%) | 63 (34%) | |
| | 2 | 296 (25%) | 35 (19%) | |
| | 3 | 301 (26%) | 49 (27%) | |
| | 4&5 | 169 (14%) | 37 (20%) | |
| Passive smoking | | | | 0.12 |
| | no | 866 (68%) | 122 (62%) | |
| | yes | 410 (32%) | 75 (38%) | |
| Puberty stage[3] | | | | 0.4 |
| | pre-pubertal | 341 (34%) | 50 (31%) | |
| | post-pubertal | 659 (66%) | 113 (69%) | |
| BMI SDS | | 1.35 (1.61) | 1.23 (1.51) | 0.3 |
| Plasma HDL-C (mmol/L) | | 1.40 (0.35) | 1.42 (0.37) | >0.9 |
| Plasma LDL-C (mmol/L) | | 2.16 (0.67) | 2.17 (0.65) | 0.9 |
| Plasma triglycerides (mmol/L) | | 0.83 (0.48) | 0.81 (0.44) | 0.7 |
| Plasma glucose (mmol/L) | | 4.96 (0.39) | 5.03 (0.36) | 0.037 |
| Plasma glucagon (pmol/L) | | 7.9 (4.3) | 7.5 (3.8) | 0.2 |
| Serum insulin (pmol/L) | | 75 (51) | 73 (38) | 0.6 |
| HOMA-IR, (mIU/L) | | 2.81 (1.98) | 2.77 (1.50) | 0.3 |
| Serum C-peptide (nmol/L) | | 0.64 (0.28) | 0.64 (0.24) | 0.5 |
| Whole blood HbA1c (mmol/mol) | | 33.08 (2.62) | 33.05 (2.66) | 0.7 |
| Obesity | | | | 0.3 |
| | no | 633 (50%) | 106 (54%) | |
| | yes | 643 (50%) | 91 (46%) | |
| Hyperglycemia | | | | 0.12 |
| | no | 1,123 (93%) | 163 (90%) | |
| | yes | 83 (6.9%) | 19 (10%) | |
| Insulin resistance | | | | >0.9 |
| | no | 985 (84%) | 148 (84%) | |
| | yes | 190 (16%) | 29 (16%) | |
| Dyslipidemia | | | | 0.5 |
| | no | 999 (82%) | 156 (84%) | |
| | yes | 220 (18%) | 29 (16%) | |
| Hypertension | | | | 0.5 |
| | no | 1,139 (90%) | 179 (92%) | |
| | yes | 122 (9.7%) | 15 (7.7%) | |

[1]Mean (SD); n (%)

[2]Wilcoxon rank sum test; Pearson's Chi-squared test

[3]Puberty stage defined as pre-pubertal (Tanner stage 1) or pubertal (Tanner stage 2-5)

***Abbrevations:** BMI SDS, body mass index standard deviation score; HDL-C, high-density lipoprotein cholesterol; LDL-C, low-density lipoprotein cholesterol; HOMA-IR, homeostasis model assessment of insulin resistance; HbA1c, glycated hemoglobin

**Table S5:** Overview over ELISA and Multiplex serology results. Missing values can be attributed to errors in the bead-counting process.

| | **ELISA** | | **Multiplex serology** |
|---|---|---|---|
| | **OD ratio** | | |
| ***H. pylori* (HP) status[a]** | **mean** | **SD** | **HP proteins[b]** |
| negative | 0.07 | 0 | 0 |
| negative | 0.04 | 0 | 1 |
| negative | 0.05 | 0 | 0 |
| negative | 0.12 | 0 | 0 |
| negative | 0.18 | 0 | 1 |
| negative | 0.19 | 0 | 1 |
| Intermediate | 0.21 | 0.01 | NA |
| Intermediate | 0.23 | 0 | 4 |
| Intermediate | 0.26 | 0 | 4 |
| Intermediate | 0.26 | 0.03 | NA |
| Intermediate | 0.3 | 0.01 | 4 |
| Intermediate | 0.35 | 0.01 | 3 |
| Intermediate | 0.36 | 0.02 | 0 |
| Intermediate | 0.36 | 0.04 | 3 |
| Intermediate | 0.36 | 0.01 | 0 |
| Intermediate | 0.38 | 0.01 | 1 |
| Intermediate | 0.41 | 0.01 | 3 |
| Intermediate | 0.43 | 0 | 1 |
| Intermediate | 0.45 | 0.01 | 2 |
| Intermediate | 0.45 | 0.01 | 2 |
| Intermediate | 0.47 | 0.06 | 4 |
| Intermediate | 0.47 | 0.03 | 1 |
| Intermediate | 0.51 | 0 | 3 |
| Intermediate | 0.52 | 0.02 | 1 |
| Intermediate | 0.56 | 0.03 | 0 |
| Intermediate | 0.57 | 0.02 | 0 |
| Intermediate | 0.6 | 0.05 | 0 |
| Intermediate | 0.6 | 0.02 | 1 |
| Intermediate | 0.63 | 0.03 | 2 |
| Intermediate | 0.67 | 0.05 | 0 |
| Intermediate | 0.68 | 0.03 | 1 |
| Intermediate | 0.69 | 0.01 | 2 |
| Intermediate | 0.7 | 0 | 2 |
| Intermediate | 0.7 | 0.01 | 0 |
| Intermediate | 0.72 | 0.04 | 0 |
| Intermediate | 0.72 | 0.02 | 1 |
| Intermediate | 0.75 | 0 | 0 |
| Intermediate | 0.76 | 0 | 1 |
| Intermediate | 0.76 | 0.01 | 2 |
| Intermediate | 0.81 | 0.01 | 1 |
| Intermediate | 0.82 | 0.02 | 0 |
| Intermediate | 0.84 | 0.04 | 3 |
| Intermediate | 0.86 | 0 | 2 |
| Intermediate | 0.93 | 0.05 | NA |
| Intermediate | 0.94 | 0.06 | 2 |
| Intermediate | 0.94 | 0.03 | NA |
| Intermediate | 0.94 | 0.01 | 6 |
| Intermediate | 0.96 | 0.01 | 1 |
| Intermediate | 0.96 | 0.04 | 2 |
| Intermediate | 0.98 | 0.01 | 2 |
| positive | 1.18 | 0.03 | NA |
| positive | 1.25 | 0.05 | 1 |
| positive | 1.34 | 0.01 | 0 |
| positive | 1.44 | 0.06 | 4 |
| positive | 1.47 | 0.03 | 4 |
| positive | 1.56 | 0.03 | 0 |
| positive | 1.63 | 0.01 | 0 |
| positive | 1.64 | 0.05 | 1 |
| positive | 2.16 | 0.06 | 0 |
| positive | 3.57 | 0.06 | 10 |

[a]positive: OD ratio ≥ 1, intermediate: 1 > OD ratio ≥ 0.2, negative: OD ratio ≤ 0.2;

[b]highlighted in bold font are *H. pylori* multiplex serology positive individuals (positive for >3 proteins)

**Table S6:** Estimated odd ratios (OR) with 95 % confidence intervals (CI) for associations of different cut-off values for *Helicobacter pylori* seropositivity as an indicator of categorical (yes/no) cardiometabolic risk factors.

| | | | | 95 % CI | | | | |
|---|---|---|---|---|---|---|---|---|
| **cutoff** | **outcome** | **n** | **OR** | **lower** | **upper** | ***p* value** | **model*** | ***H. pylori* seropositivity** |
| 0.9 | Obesity | 1703 | 1.061 | 0.820 | 1.374 | 0.655 | 1 | 16.4 |
| 0.9 | Obesity | 1531 | 0.925 | 0.682 | 1.254 | 0.616 | 2 | 16.4 |
| 0.9 | Obesity | 1209 | 1.023 | 0.724 | 1.446 | 0.895 | 3 | 16.4 |
| 0.9 | Hyperglycemia | 1596 | 1.880 | 1.219 | 2.838 | 0.003 | 1 | 16.4 |
| 0.9 | Hyperglycemia | 1436 | 1.819 | 1.135 | 2.844 | 0.010 | 2 | 16.4 |
| 0.9 | Hyperglycemia | 1151 | 2.231 | 1.346 | 3.615 | 0.001 | 3 | 16.4 |
| 0.9 | Hypertension | 1681 | 1.020 | 0.644 | 1.566 | 0.931 | 1 | 16.4 |
| 0.9 | Hypertension | 1514 | 0.830 | 0.491 | 1.345 | 0.467 | 2 | 16.4 |
| 0.9 | Hypertension | 1199 | 0.951 | 0.547 | 1.583 | 0.851 | 3 | 16.4 |
| 0.9 | Dyslipidemia | 1615 | 0.855 | 0.582 | 1.237 | 0.415 | 1 | 16.4 |
| 0.9 | Dyslipidemia | 1455 | 0.879 | 0.579 | 1.310 | 0.536 | 2 | 16.4 |
| 0.9 | Dyslipidemia | 1164 | 0.785 | 0.488 | 1.231 | 0.303 | 3 | 16.4 |
| 0.9 | Insulin resistance | 1558 | 1.302 | 0.898 | 1.869 | 0.158 | 1 | 16.4 |
| 0.9 | Insulin resistance | 1404 | 1.189 | 0.789 | 1.769 | 0.400 | 2 | 16.4 |
| 0.9 | Insulin resistance | 1122 | 1.430 | 0.908 | 2.224 | 0.116 | 3 | 16.4 |
| 0.8 | Obesity | 1703 | 1.023 | 0.798 | 1.313 | 0.856 | 1 | 18.1 |
| 0.8 | Obesity | 1531 | 0.909 | 0.678 | 1.218 | 0.522 | 2 | 18.1 |
| 0.8 | Obesity | 1209 | 0.920 | 0.658 | 1.284 | 0.623 | 3 | 18.1 |
| 0.8 | Hyperglycemia | 1596 | 1.749 | 1.141 | 2.627 | 0.008 | 1 | 18.1 |
| 0.8 | Hyperglycemia | 1436 | 1.692 | 1.063 | 2.631 | 0.022 | 2 | 18.1 |
| 0.8 | Hyperglycemia | 1151 | 2.002 | 1.211 | 3.233 | 0.005 | 3 | 18.1 |
| 0.8 | Hypertension | 1681 | 0.999 | 0.640 | 1.517 | 0.995 | 1 | 18.1 |
| 0.8 | Hypertension | 1514 | 0.826 | 0.499 | 1.317 | 0.440 | 2 | 18.1 |
| 0.8 | Hypertension | 1199 | 0.993 | 0.584 | 1.626 | 0.977 | 3 | 18.1 |
| 0.8 | Dyslipidemia | 1615 | 0.919 | 0.637 | 1.310 | 0.647 | 1 | 18.1 |
| 0.8 | Dyslipidemia | 1455 | 0.930 | 0.624 | 1.364 | 0.714 | 2 | 18.1 |
| 0.8 | Dyslipidemia | 1164 | 0.802 | 0.505 | 1.243 | 0.335 | 3 | 18.1 |
| 0.8 | Insulin resistance | 1558 | 1.284 | 0.895 | 1.825 | 0.169 | 1 | 18.1 |
| 0.8 | Insulin resistance | 1404 | 1.175 | 0.789 | 1.729 | 0.418 | 2 | 18.1 |
| 0.8 | Insulin resistance | 1122 | 1.348 | 0.861 | 2.083 | 0.184 | 3 | 18.1 |
| 0.7 | Obesity | 1703 | 0.994 | 0.785 | 1.260 | 0.962 | 1 | 20.6 |
| 0.7 | Obesity | 1531 | 0.905 | 0.685 | 1.194 | 0.481 | 2 | 20.6 |
| 0.7 | Obesity | 1209 | 0.890 | 0.647 | 1.222 | 0.471 | 3 | 20.6 |
| 0.7 | Hyperglycemia | 1596 | 1.623 | 1.074 | 2.411 | 0.019 | 1 | 20.6 |
| 0.7 | Hyperglycemia | 1436 | 1.599 | 1.022 | 2.454 | 0.035 | 2 | 20.6 |
| 0.7 | Hyperglycemia | 1151 | 1.960 | 1.208 | 3.123 | 0.005 | 3 | 20.6 |
| 0.7 | Hypertension | 1681 | 1.058 | 0.699 | 1.568 | 0.784 | 1 | 20.6 |
| 0.7 | Hypertension | 1514 | 0.941 | 0.594 | 1.448 | 0.787 | 2 | 20.6 |
| 0.7 | Hypertension | 1199 | 1.155 | 0.711 | 1.827 | 0.549 | 3 | 20.6 |
| 0.7 | Dyslipidemia | 1615 | 1.053 | 0.749 | 1.464 | 0.764 | 1 | 20.6 |
| 0.7 | Dyslipidemia | 1455 | 1.118 | 0.776 | 1.594 | 0.542 | 2 | 20.6 |
| 0.7 | Dyslipidemia | 1164 | 1.010 | 0.665 | 1.512 | 0.962 | 3 | 20.6 |
| 0.7 | Insulin resistance | 1558 | 1.157 | 0.819 | 1.622 | 0.401 | 1 | 20.6 |
| 0.7 | Insulin resistance | 1404 | 1.048 | 0.716 | 1.519 | 0.805 | 2 | 20.6 |
| 0.7 | Insulin resistance | 1122 | 1.252 | 0.815 | 1.899 | 0.297 | 3 | 20.6 |

*model 1 adjusted for age, sex, BMI SDS (except for the outcome "Obesity")
model 2: model 1 + additional adjustment for socioeconomic status
model 3: model 1 + model 2 + additional adjustment for puberty stage

**Table S7**: Standardized coefficient (beta) estimates with 95 % confidence intervals (CI) for associations of different cut-off values for *Helicobacter pylori* seropositivity as an indicator of continuous cardiometabolic risk factors. Outcome variables were log10-transformed and z-scored, except for BMI SDS, bodyfat % SDS and WtHR SDS.

| | | | | 95 % CI | | | |
|---|---|---|---|---|---|---|---|
| **cutoff** | **outcome** | **n** | **beta** | **lower** | **upper** | ***p* value** | **model** |
| 0.9 | BMI SDS | 1703 | 0.0923 | -0.1108 | 0.2955 | 0.3732 | 1 |
| 0.9 | BMI SDS | 1531 | 0.0003 | -0.1920 | 0.1926 | 0.9972 | 2 |
| 0.9 | BMI SDS | 1209 | 0.0499 | -0.1595 | 0.2593 | 0.6407 | 3 |
| 0.9 | Body fat % SDS | 698 | -0.0213 | -0.2039 | 0.1613 | 0.8195 | 1 |
| 0.9 | Body fat % SDS | 609 | 0.0101 | -0.1647 | 0.1848 | 0.9102 | 2 |
| 0.9 | Body fat % SDS | 474 | -0.0392 | -0.2342 | 0.1557 | 0.6934 | 3 |
| 0.9 | WtHR SDS | 1610 | 0.0504 | -0.1221 | 0.2229 | 0.5671 | 1 |
| 0.9 | WtHR SDS | 1452 | -0.0320 | -0.1964 | 0.1323 | 0.7028 | 2 |
| 0.9 | WtHR SDS | 1155 | -0.0125 | -0.1938 | 0.1687 | 0.8922 | 3 |
| 0.9 | Glucose | 1602 | 0.1619 | 0.0310 | 0.2929 | 0.0155 | 1 |
| 0.9 | Glucose | 1441 | 0.1285 | -0.0105 | 0.2675 | 0.0703 | 2 |
| 0.9 | Glucose | 1156 | 0.1913 | 0.0381 | 0.3444 | 0.0145 | 3 |
| 0.9 | Insulin | 1626 | 0.0695 | -0.0374 | 0.1763 | 0.2030 | 1 |
| 0.9 | Insulin | 1462 | 0.0423 | -0.0688 | 0.1534 | 0.4559 | 2 |
| 0.9 | Insulin | 1171 | 0.0781 | -0.0439 | 0.2001 | 0.2099 | 3 |
| 0.9 | HOMA | 1595 | 0.0971 | -0.0144 | 0.2086 | 0.0881 | 1 |
| 0.9 | HOMA | 1434 | 0.0587 | -0.0584 | 0.1757 | 0.3259 | 2 |
| 0.9 | HOMA | 1150 | 0.1002 | -0.0279 | 0.2283 | 0.1256 | 3 |
| 0.9 | HbA1c | 1613 | 0.0043 | -0.0004 | 0.0091 | 0.0755 | 1 |
| 0.9 | HbA1c | 1453 | 0.0029 | -0.0021 | 0.0078 | 0.2560 | 2 |
| 0.9 | HbA1c | 1164 | 0.0652 | -0.0853 | 0.2157 | 0.3957 | 3 |
| 0.9 | C-peptide | 1582 | 0.0647 | -0.0374 | 0.1669 | 0.2142 | 1 |
| 0.9 | C-peptide | 1423 | 0.0590 | -0.0475 | 0.1655 | 0.2777 | 2 |
| 0.9 | C-peptide | 1150 | 0.0969 | -0.0172 | 0.2109 | 0.0962 | 3 |
| 0.8 | BMI SDS | 1703 | 0.0554 | -0.1406 | 0.2514 | 0.5798 | 1 |
| 0.8 | BMI SDS | 1531 | -0.0280 | -0.2132 | 0.1572 | 0.7671 | 2 |
| 0.8 | BMI SDS | 1209 | -0.0249 | -0.2273 | 0.1776 | 0.8100 | 3 |
| 0.8 | Body fat % SDS | 698 | -0.0213 | -0.2039 | 0.1613 | 0.8195 | 1 |
| 0.8 | Body fat % SDS | 609 | 0.0174 | -0.1529 | 0.1876 | 0.8416 | 2 |
| 0.8 | Body fat % SDS | 474 | -0.0319 | -0.2234 | 0.1596 | 0.7442 | 3 |
| 0.8 | WtHR SDS | 1610 | 0.0184 | -0.1478 | 0.1845 | 0.8285 | 1 |
| 0.8 | WtHR SDS | 1452 | -0.0542 | -0.2122 | 0.1038 | 0.5018 | 2 |
| 0.8 | WtHR SDS | 1155 | -0.0679 | -0.2429 | 0.1071 | 0.4473 | 3 |
| 0.8 | Glucose | 1602 | 0.1811 | 0.0549 | 0.3074 | 0.0050 | 1 |
| 0.8 | Glucose | 1441 | 0.1563 | 0.0225 | 0.2901 | 0.0222 | 2 |
| 0.8 | Glucose | 1156 | 0.2083 | 0.0605 | 0.3562 | 0.0058 | 3 |
| 0.8 | Insulin | 1626 | 0.0705 | -0.0327 | 0.1736 | 0.1810 | 1 |
| 0.8 | Insulin | 1462 | 0.0522 | -0.0549 | 0.1594 | 0.3393 | 2 |
| 0.8 | Insulin | 1171 | 0.0735 | -0.0445 | 0.1915 | 0.2224 | 3 |
| 0.8 | HOMA | 1595 | 0.1008 | -0.0067 | 0.2083 | 0.0664 | 1 |
| 0.8 | HOMA | 1434 | 0.0725 | -0.0402 | 0.1853 | 0.2074 | 2 |
| 0.8 | HOMA | 1150 | 0.0978 | -0.0260 | 0.2216 | 0.1217 | 3 |
| 0.8 | HbA1c | 1613 | 0.0041 | -0.0005 | 0.0087 | 0.0774 | 1 |
| 0.8 | HbA1c | 1453 | 0.0030 | -0.0017 | 0.0078 | 0.2087 | 2 |
| 0.8 | HbA1c | 1164 | 0.0719 | -0.0735 | 0.2173 | 0.3328 | 3 |
| 0.8 | C-peptide | 1582 | 0.0741 | -0.0243 | 0.1725 | 0.1400 | 1 |
| 0.8 | C-peptide | 1423 | 0.0745 | -0.0279 | 0.1770 | 0.1542 | 2 |
| 0.8 | C-peptide | 1150 | 0.1014 | -0.0089 | 0.2117 | 0.0718 | 3 |
| 0.7 | BMI SDS | 1703 | 0.0544 | -0.1317 | 0.2406 | 0.5664 | 1 |
| 0.7 | BMI SDS | 1531 | -0.0016 | -0.1772 | 0.1740 | 0.9858 | 2 |
| 0.7 | BMI SDS | 1209 | -0.0133 | -0.2057 | 0.1791 | 0.8924 | 3 |
| 0.7 | Body fat % SDS | 698 | -0.0213 | -0.2039 | 0.1613 | 0.8195 | 1 |
| 0.7 | Body fat % SDS | 609 | 0.0135 | -0.1480 | 0.1751 | 0.8696 | 2 |
| 0.7 | Body fat % SDS | 474 | -0.0410 | -0.2224 | 0.1404 | 0.6580 | 3 |
| 0.7 | WtHR SDS | 1610 | 0.0289 | -0.1291 | 0.1869 | 0.7202 | 1 |
| 0.7 | WtHR SDS | 1452 | -0.0296 | -0.1798 | 0.1206 | 0.6996 | 2 |
| 0.7 | WtHR SDS | 1155 | -0.0536 | -0.2201 | 0.1129 | 0.5279 | 3 |
| 0.7 | Glucose | 1602 | 0.1294 | 0.0098 | 0.2491 | 0.0341 | 1 |
| 0.7 | Glucose | 1441 | 0.1049 | -0.0215 | 0.2314 | 0.1042 | 2 |
| 0.7 | Glucose | 1156 | 0.1713 | 0.0312 | 0.3114 | 0.0167 | 3 |
| 0.7 | Insulin | 1626 | 0.0472 | -0.0507 | 0.1450 | 0.3447 | 1 |
| 0.7 | Insulin | 1462 | 0.0278 | -0.0735 | 0.1292 | 0.5904 | 2 |
| 0.7 | Insulin | 1171 | 0.0630 | -0.0489 | 0.1749 | 0.2698 | 3 |
| 0.7 | HOMA | 1595 | 0.0715 | -0.0303 | 0.1733 | 0.1687 | 1 |
| 0.7 | HOMA | 1434 | 0.0421 | -0.0644 | 0.1486 | 0.4385 | 2 |
| 0.7 | HOMA | 1150 | 0.0837 | -0.0334 | 0.2009 | 0.1615 | 3 |
| 0.7 | HbA1c | 1613 | 0.0010 | -0.0033 | 0.0054 | 0.6368 | 1 |
| 0.7 | HbA1c | 1453 | 0.0006 | -0.0039 | 0.0051 | 0.7821 | 2 |
| 0.7 | HbA1c | 1164 | 0.0030 | -0.1347 | 0.1408 | 0.9657 | 3 |
| 0.7 | C-peptide | 1582 | 0.0530 | -0.0403 | 0.1464 | 0.2657 | 1 |
| 0.7 | C-peptide | 1423 | 0.0505 | -0.0466 | 0.1475 | 0.3082 | 2 |
| 0.7 | C-peptide | 1150 | 0.0837 | -0.0210 | 0.1885 | 0.1175 | 3 |

*model 1 adjusted for age, sex, BMI SDS (except for the outcome "BMI SDS", "Body fat % SDS" and "WtHR SDS")
model 2: model 1 + additional adjustment for socioeconomic status
model 3: model 1 + model 2 + additional adjustment for puberty stage

**Table S8:** Estimated odd ratios (OR) with 95 % confidence intervals (CI) for associations of *Helicobacter pylori* seropositivity as an indicator of categorical (yes/no) cardiometabolic risk factors. 71 subjects with non-European genetic ethnicity were excluded.

| | Model 1[a] | | | Model 2[b] | | | Model 3[c] | | |
|---|---|---|---|---|---|---|---|---|---|
| | **n*** | **OR (95% CI)** | **p value** | **n*** | **OR (95% CI)** | **p value** | **n*** | **OR (95% CI)** | **p value** |
| Obesity | 1632 | 0.93 (0.70;1.23) | 0.61 | 1484 | 0.83 (0.60;1.15) | 0.28 | 1170 | 0.94 (0.65;1.36) | 0.75 |
| Hyperglycemia | 1529 | 2.21 (1.40;3.42) | 0.0005 | 1391 | 2.11 (1.28;3.37) | 0.002 | 1114 | 2.49 (1.46;4.14) | 0.001 |
| Hypertension | 1614 | 0.97 (0.57;1.56) | 0.89 | 1468 | 0.79 (0.44;1.35) | 0.41 | 1161 | 0.84 (0.45;1.47) | 0.55 |
| Dyslipidemia | 1547 | 0.86 (0.56;1.29) | 0.47 | 1409 | 0.83 (0.52;1.29) | 0.42 | 1126 | 0.70 (0.41;1.15) | 0.17 |
| Insulin resistance | 1492 | 1.34 (0.89;2.00) | 0.16 | 1359 | 1.25 (0.80;1.93) | 0.31 | 1085 | 1.49 (0.92;2.39) | 0.10 |

[a]adjusted for age, sex, BMI SDS (except for the outcome "obesity")
[b]additional adjustment for socioeconomic status
[c]additional adjustment for puberty stage
*sample size after removal of missing values

**Table S9:** Standardized coefficient (beta) estimates with 95 % confidence intervals (CI) for associations of *Helicobacter pylori* seropositivity as an indicator of continuous cardiometabolic risk factors. Outcome variables were log10-transformed and z-scored except for body mass index (BMI) standard deviation score (SDS), bodyfat % SDS and waist to height ratio (WtHR) SDS. 71 subjects with non-European genetic ethnicity were excluded.

| | Model 1[a] | | | Model 2[b] | | | Model 3[c] | | |
|---|---|---|---|---|---|---|---|---|---|
| | **n*** | **beta (95% CI)** | ***p*** | **n*** | **beta (95% CI)** | ***p*** | **n*** | **beta (95% CI)** | ***p*** |
| BMI SDS | 1632 | -0.02 (-0.24;0.20) | 0.88 | 1484 | -0.06 (-0.27;0.14) | 0.56 | 1170 | 0.01 (-0.21;0.24) | 0.92 |
| Body fat (%) SDS | 649 | -0.05 (-0.24;0.14) | 0.61 | 573 | 0.00 (-0.19;0.20) | 0.98 | 442 | -0.03 (-0.24;0.18) | 0.79 |
| WtHR SDS | 1546 | -0.063 (-0.25;0.12) | 0.51 | 1408 | -0.105 (-0.28;0.07) | 0.25 | 1118 | -0.06 (-0.25;0.13) | 0.55 |
| Glucose | 1535 | 0.22 (0.08;0.37) | 0.002 | 1396 | 0.18 (0.03;0.33) | 0.017 | 1119 | 0.22 (0.06;0.38) | 0.009 |
| Insulin | 1557 | 0.08 (-0.03;0.20) | 0.16 | 1416 | 0.06 (-0.06;0.18) | 0.34 | 1133 | 0.09 (-0.04;0.22) | 0.18 |
| HOMA-IR | 1528 | 0.12 (-0.01;0.24) | 0.06 | 1389 | 0.08 (-0.05;0.21) | 0.22 | 1113 | 0.11 (-0.03;0.25) | 0.11 |
| HbA1c | 1545 | 0.004 (-0.001;0.01) | 0.10 | 1407 | 0.003 (-0.003;0.008) | 0.32 | 1126 | 0.07 (-0.10;0.23) | 0.42 |
| C-peptide | 1515 | 0.064 (-0.0474;0.17) | 0.26 | 1379 | 0.06 (-0.06;0.18) | 0.31 | 1114 | 0.11 (-0.02;0.23) | 0.09 |

[a]adjusted for age, sex, BMI SDS (except for the outcome "BMI SDS", "Bodyfat % SDS" and "WtHR SDS")
[b]additional adjustment for socioeconomic status
[c]additional adjustment for puberty stage
*sample size after removal of missing values

**Table S10:** Estimated odd ratios (OR) with 95 % confidence intervals (CI) for associations of *Helicobacter pylori* seropositivity as an indicator of categorical (yes/no) cardiometabolic risk factors. 71 subjects with non-European genetic ethnicity and 159 subjects with self-reported non-Danish ethnicity were excluded.

| | Model 1[a] | | | Model 2[b] | | | Model 3[c] | | |
|---|---|---|---|---|---|---|---|---|---|
| | **n*** | **OR (95% CI)** | **p value** | **n*** | **OR (95% CI)** | **p value** | **n*** | **OR (95% CI)** | **p value** |
| Obesity | 1473 | 0.87 (0.64;1.18) | 0.36 | 1354 | 0.75 (0.53;1.06) | 0.10 | 1073 | 0.83 (0.56;1.22) | 0.35 |
| Hyperglycemia | 1388 | 1.64 (0.94;2.74) | 0.07 | 1275 | 1.58 (0.87;2.72) | 0.12 | 1025 | 2.01 (1.09;3.55) | 0.02 |
| Hypertension | 1455 | 0.85 (0.46;1.47) | 0.58 | 1338 | 0.80 (0.41;1.43) | 0.47 | 1064 | 0.81 (0.40;1.50) | 0.53 |
| Dyslipidemia | 1404 | 0.92 (0.57;1.43) | 0.71 | 1291 | 0.93 (0.56;1.49) | 0.76 | 1036 | 0.78 (0.44;1.32) | 0.36 |
| Insulin resistance | 1352 | 1.15 (0.72;1.81) | 0.54 | 1244 | 1.10 (0.67;1.77) | 0.69 | 997 | 1.26 (0.73;2.11) | 0.39 |

[a]adjusted for age, sex, BMI SDS (except for the outcome "obesity")
[b]additional adjustment for socioeconomic status
[c]additional adjustment for puberty stage
*sample size after removal of missing values

**Table S11:** Standardized coefficient (beta) estimates with 95 % confidence intervals (CI) for associations of *Helicobacter pylori* seropositivity as an indicator of continuous cardiometabolic risk factors. Outcome variables were log10-transformed and z-scored except for body mass index (BMI) standard deviation score (SDS), bodyfat % SDS and waist to height ratio (WtHR) SDS. 71 subjects with non-European genetic ethnicity and 159 subjects with self-reported non-Danish ethnicity were excluded.

| | Model 1[a] | | | Model 2[b] | | | Model 3[c] | | |
|---|---|---|---|---|---|---|---|---|---|
| | **n*** | **beta (95% CI)** | ***p*** | **n*** | **beta (95% CI)** | ***p*** | **n*** | **beta (95% CI)** | ***p*** |
| BMI SDS | 1473 | -0.08 (-0.32;0.16) | 0.51 | 1354 | -0.15 (-0.37;0.07) | 0.19 | 1073 | -0.07 (-0.31;0.17) | 0.56 |
| Body fat (%) SDS | 580 | -0.08 (-0.30;0.13) | 0.44 | 514 | -0.02 (-0.23;0.19) | 0.86 | 398 | -0.04 (-0.27;0.19) | 0.71 |
| WtHR SDS | 1396 | -0.095 (-0.30;0.11) | 0.35 | 1286 | -0.153 (-0.34;0.04) | 0.11 | 1024 | -0.10 (-0.31;0.10) | 0.32 |
| Glucose | 1393 | 0.19 (0.03;0.34) | 0.017 | 1279 | 0.17 (0.01;0.33) | 0.04 | 1029 | 0.22 (0.05;0.39) | 0.013 |
| Insulin | 1410 | 0.02 (-0.11;0.15) | 0.76 | 1296 | 0.01 (-0.13;0.14) | 0.93 | 1041 | 0.03 (-0.11;0.17) | 0.70 |
| HOMA-IR | 1387 | 0.06 (-0.07;0.19) | 0.35 | 1273 | 0.05 (-0.09;0.18) | 0.50 | 1024 | 0.08 (-0.07;0.22) | 0.31 |
| HbA1c | 1403 | 0.0001 (-0.005;0.01) | 0.98 | 1290 | -0.001 (-0.006;0.005) | 0.79 | 1037 | -0.02 (-0.19;0.16) | 0.85 |
| C-peptide | 1373 | 0.027 (-0.0932;0.15) | 0.66 | 1264 | 0.02 (-0.11;0.14) | 0.79 | 1025 | 0.05 (-0.08;0.18) | 0.42 |

[a]adjusted for age, sex, BMI SDS (except for the outcome "BMI SDS", "Bodyfat % SDS" and "WtHR SDS")
[b]additional adjustment for socioeconomic status
[c]additional adjustment for puberty stage
*sample size after removal of missing values

**Table S12:** Multiplex serology *Helicobacter pylori* antigens and antigen-specific cut-offs at 1:100 serum dilution

| Name | Systematic name | Acc. No[a] | Selected amino acids | Cut-off [MFI] |
|---|---|---|---|---|
| ***H. pylori*** | | | | |
| GroEl | HP0010 | AM_997163 | 1-547 | 400 |
| UreA | HP0073 | NP_206873 | 1-238 | 1250 |
| HP0231 | HP0231 | NP_207029 | 1-265 | 100 |
| NapA | HP0243 | NP_207041 | 1-144 | 280 |
| HP0305 | HP0305 | NP_207103 | 1-184 | 150 |
| HpaA | HP0410 | NP_207208 | 1-249 | 650 |
| CagA N-Terminus | HP0547 | NP_207343 | 1-650 | 1000 |
| HyuA N-Terminus | HP0695 | NP_207208 | 1-220 | 600 |
| VacA C-Terminus | HP0887 | NP_207680 | 328-1,008 | 600 |
| HcpC | HP1098 | NP_207889 | 1-290 | 200 |
| Cad | HP1104 | NP_207895 | 1-348 | 270 |
| HP1564 | HP1564 | NP_208355 | 1-271 | 1750 |

[a]NCBI reference sequence
